# The Role of Social Vulnerability: Temporal Patterns of County-Level Health Disparities in the State of Indiana

**DOI:** 10.64898/2026.08.05.26359801

**Authors:** Kelli Wang, Precious Olaniyan, Plamena Powla, Felix Pabon Rodriguez

## Abstract

Indiana still faces significant health challenges, ranking among the least healthy U.S. states due to high obesity rates, mental health issues, and other chronic conditions. These disparities are closely linked to inequities in healthcare access, which are largely shaped by social determinants of health. Using data from the Social Vulnerability Index and County Health Rankings and Roadmaps, this study analyzes trends in obesity, mental health, and premature death across Indiana counties before, during, and after the COVID-19 pandemic. Descriptive statistics, correlation analyses, and Negative Binomial regression models were used to evaluate county-level disparities. In 2018, higher rates of uninsured, obese, and physically inactive populations were associated with increased premature death. In 2020, diabetes, smoking, and alcohol consumption were significant factors. By 2022, unemployment, education, obesity, insurance, exercise access, and mental health provider availability were associated with premature death. Findings indicate that socially vulnerable counties experienced amplified health impacts, with obesity rising most sharply where exercise infrastructure was limited and poor mental health days increasing across all counties. These results highlight persistent service gaps and the critical need for targeted investments in recreational infrastructure and mental healthcare. Future research should examine policy influences and causal relationships to inform equity-focused interventions.

## Introduction

Mental health and obesity are two increasing public health issues, nationally, and in the state of Indiana.^1^ Indiana records from the County Health Rankings and Roadmaps (CHR&R) show that the percentage of adult obesity increased from 32% in 2018 to 35% 2022, while mental health issues, defined as poor mental health days, increased from 4.3 to 4.8 days. Both conditions show estimates above the United States (US) level.^2^ When looking at premature death, defined as the years of potential life lost before age 75 per 100,000 population (denoted by YPLL-75), Indiana also shows a higher measure compared to the US level (7,800 vs 6,700 in 2018; and 8,600 vs 7,300 in 2022). The percentage of adult smoking and adults that are physically inactive are also higher in Indiana compared to the US.

Social and economic vulnerabilities have significantly shaped healthcare access and health outcomes in Indiana both prior to and throughout the COVID-19 pandemic, and their effects persist in its aftermath. Research on social determinants of health highlights that income, education, and wealth are powerful predictors of health, often exerting more influence than direct medical care.^3^ Even when healthcare access is equalized, disparities in outcomes remain, particularly among individuals in lower socioeconomic positions. These long-standing structural inequalities create a gradient where poorer Hoosiers (Indiana residents) experience reduced life expectancy and diminished well-being.

Prior to the pandemic, obesity was already a major public health crisis in Indiana, disproportionately affecting populations marginalized by poverty, race, and gender.^4^ Studies show that Black women and those living in urban poverty in Indianapolis had limited access to nutritious food,^4^ relied heavily on fast food, and lacked resources for exercise, all contributing to a persistently high average body mass index (BMI). The 2019 obesity report further confirms these trends,^5^ attributing rising obesity to systemic issues like poor diet, limited physical activity, and socioeconomic constraints, with significant economic consequences for the state. Obesity rates continued to increase during and after the pandemic, with the rate remaining significantly higher than pre-pandemic obesity levels. For example, obesity rates for children spiked during the pandemic, peaking in 2021. By collecting data from over 400,000 patients in various Indiana counties, researchers found that the pandemic-era surge in obesity rates has not significantly reversed.^6^

The pandemic also had a significant impact on mental health and substance abuse. In 2021, the US recorded its highest number of deaths by drug overdose, which mainly affected people of color.^7^ Opioid overdoses were a big driver for the increase in drug overdose, accounting for 75% of drug overdoses in 2021.^7^ In Indiana, suicide rates significantly increased during the pandemic, and the state rate was higher than the national rate.^7^ About 31% of Indiana’s need for mental health professionals are met, which is higher than the national percentage, but rural areas are often unable to access mental health resources in a timely manner.^7^ Untreated mental health illnesses will eventually become an economic burden to Indiana, with an estimated burden of $4.2 billion.^8^ This cost came primarily from indirect losses such as unemployment and reduced productivity.^8^ These figures reflect not only inadequate care but also the cumulative impact of decades of social and economic disadvantage.

Achieving health equity requires dismantling the entrenched barriers of poverty, discrimination, and unequal opportunity. This goes for Indiana as well, achieving this equity involves not only expanding healthcare access, but also empowering marginalized communities to help design and implement solutions tailored to their lived experiences. Only by assessing these root causes can Indiana hope to improve health outcomes across all counties, especially for its most vulnerable residents. This project aims to aid in exploring potential influencing factors of health disparities by analyzing health trends across Indiana counties so that long-term solutions can be developed and implemented.

## Methods

This cross-sectional study uses two datasets: (1) The County Health Rankings & Roadmaps (CHR&R),^2^ obtained from the University of Wisconsin Population Health Institute’s public website, and (2) The Social Vulnerability Index (SVI) from the Centers for Disease Control and Prevention (CDC).^9^ Since the focus of the study was the period before, during, and after the COVID-19 pandemic, datasets for the years 2018, 2020, and 2022 were selected. Year-specific analyses were employed. The primary outcome of the study is premature death, defined within CHR&R as years of potential life lost before age 75 per 100,000 population, and denoted by YPPL-75. Higher YPLL-75 value indicates a greater number of years lost due to premature deaths, suggesting a higher burden of premature mortality. Secondary outcomes of interest are the number of poor mental and physical health days.

We considered covariates from different domains of the social determinants of health (SDOH) framework. Sociodemographic variables included in the social vulnerability index (SVI) groups, income, insurance, education, and employment status. Behavioral variables include smoking, drinking, and exercise practices. Access to healthcare and additional services was assessed by considering the number of mental health providers, number of physicians, number of dentists, access to exercise opportunities, and food environment index. Regarding comorbidity conditions, we looked at the prevalence of obesity and diabetes. The SVI indicates the relative vulnerability of every Indiana county. SVI ranks the counties on 15 social factors, including unemployment, minority status, and disability, and further groups them into four related themes. The overall SVI ranks as well as the theme ranks are based on percentiles. Percentile ranking values range from 0 to 1, with higher values indicating greater vulnerability. For this study, we used the SVI overall ranks to define four categories (SVI groups) based on quartiles (0.00-0.25, 0.25-0.50, 0.50-0.75, 0.75-1.00) to group counties on vulnerability groups, where higher SVI indicates more social vulnerability, similarly, falling in the 3^rd^ or 4^th^ categories.

To evaluate the best distributional assumption for the primary outcome, we performed a comparative assessment between Poisson and Negative Binomial (NB) distribution. We compared the sample means and variances and investigated whether there was a signal of overdispersion (variance > mean). This was confirmed by overdispersion tests and assessments of model fits (Akaike Information Criterion and Bayesian Information Criterion, also denoted by AIC and BIC, respectively).^10^ An overdispersion test (p < 0.05) and estimates of AIC/BIC supported the choice of NB over the Poisson regression model.^10^ The next step was looking at a variable selection approach along with importance of the variable from established literature. We looked at bivariate associations between candidate predictors and the outcome. All variables entering the model were statistically associated with the outcome at the 10% significance level, this in addition to scientific knowledge and literature support. Premature death as the outcome was modeled via a Negative Binomial Regression Model^10^ for each year, accounting for overdispersion and including predictors previously identified to be associated with the outcome. This non-spatial model was considered against a spatial autocorrelation test, using the Global Moran’s I test.^11^ Results from this test, for each year, suggested no significant spatial dependence (p > 0.05). Therefore, the remaining of the manuscript is constructed based on results from the non-spatial NB regression model.

The data were obtained from the public websites and analyzed using regression models in R statistical software version 4.5.1. Figures and tables were prepared using the R packages *ggplot2*,^*12*^ and *gtsummary*,^*13*^ respectively. Data manipulation and processing were performed using the R package *dplyr*. ^*14*^

## Results and Discussion

### Results

In this study, we were interested in studying the associations between different potential influencing factors (social, clinical, behavioral, environmental, access to health care or services, contextual, etc.) and health outcomes (obesity rates, mental health, and premature death). We employed correlation analyses, temporal trends stratified by social vulnerability groups, and regression analyses.

Poor mental health days increased by an average of about 1 day from 2018 to 2022 across all counties and SVI quartiles (SVI-Qs) (**Figure 1**). From 2018 to 2022, premature death rates dramatically rose in the SVI-Q1 by nearly 33%; SVI-Q2 increased by about 1%; and 4^th^ SVI-Q increased by approximately 5%, while the SVI-Q3 saw a decrease of about 3%. However, from 2020 to 2022, all SVI-Qs experienced increases in premature death rates. Poor physical health days increased steadily from 2018 to 2022 in every SVI-Q, with a more pronounced rise for SVI-Q1 of about 1 day.

**Figure 1.**
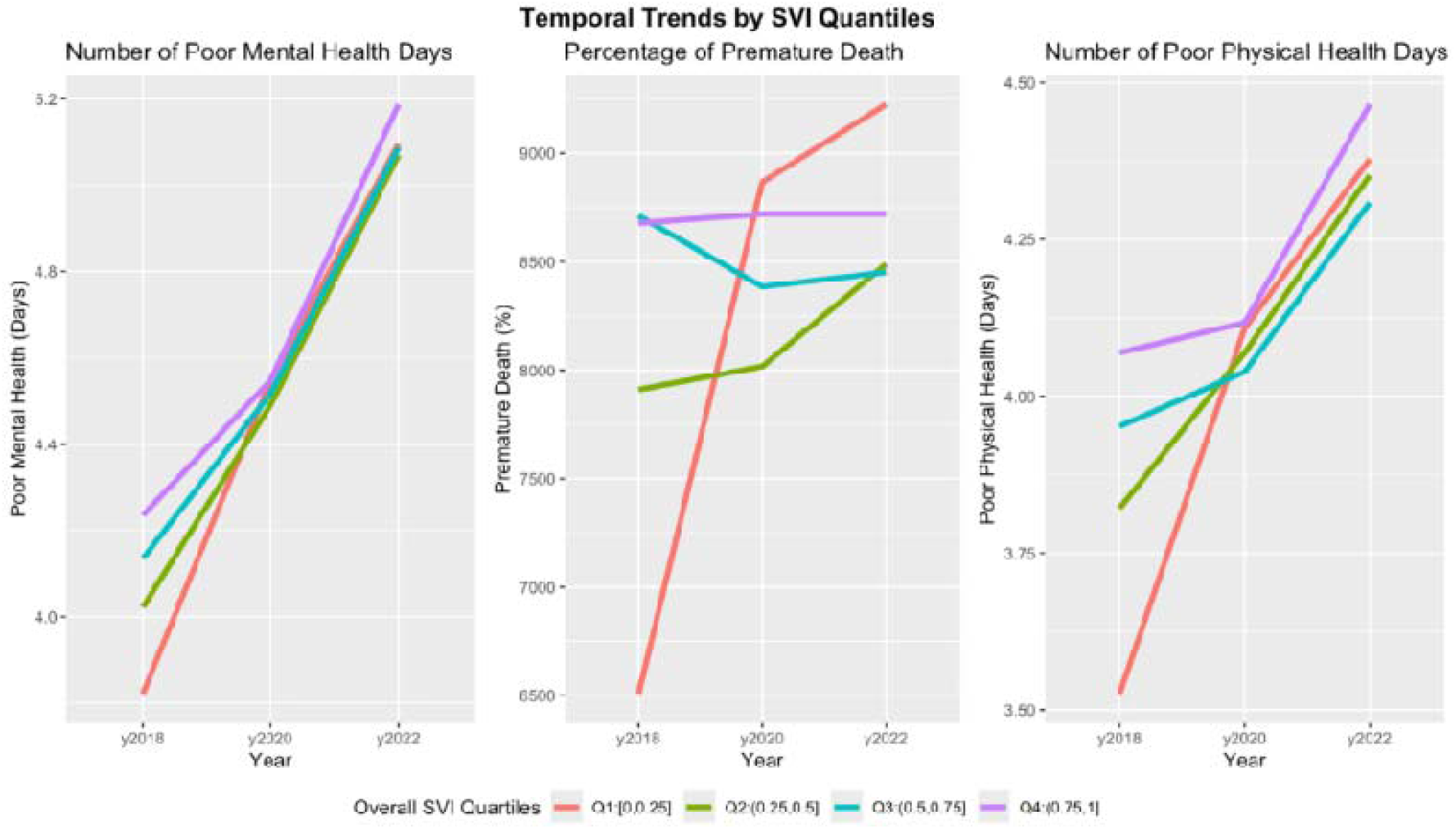
Temporal trends of health outcomes of interests by social vulnerability groups.

From 2018 to 2022, obesity rates increased across all counties in each SVI-Q (**Figure 2**). From 2018 to 2022, SVI-Q1 increased by ~5%; SVI-Q2 and 3 increased by ~2.5%; and SVI-Q4 slightly decreased in 2020 but increased overall by ~2.5% as well. During the same period, the percentage of people with diabetes decreased in all counties, regardless of SVI-Q: Q1 had a net decrease of ~0.5%; Q2 had a net decrease of ~2%; Q3 had a net decrease of ~1.5%; and Q4 had a net decrease of ~1%. In contrast, smoking rates increased across all counties in each SVI-Q between 2018 and 2022, with a notable increase in Q1 of about 4%. Alcohol consumption declined from 2018 to 2020 but reversed course and increased from 2020 to 2022 across all SVI-Qs.

**Figure 2.**
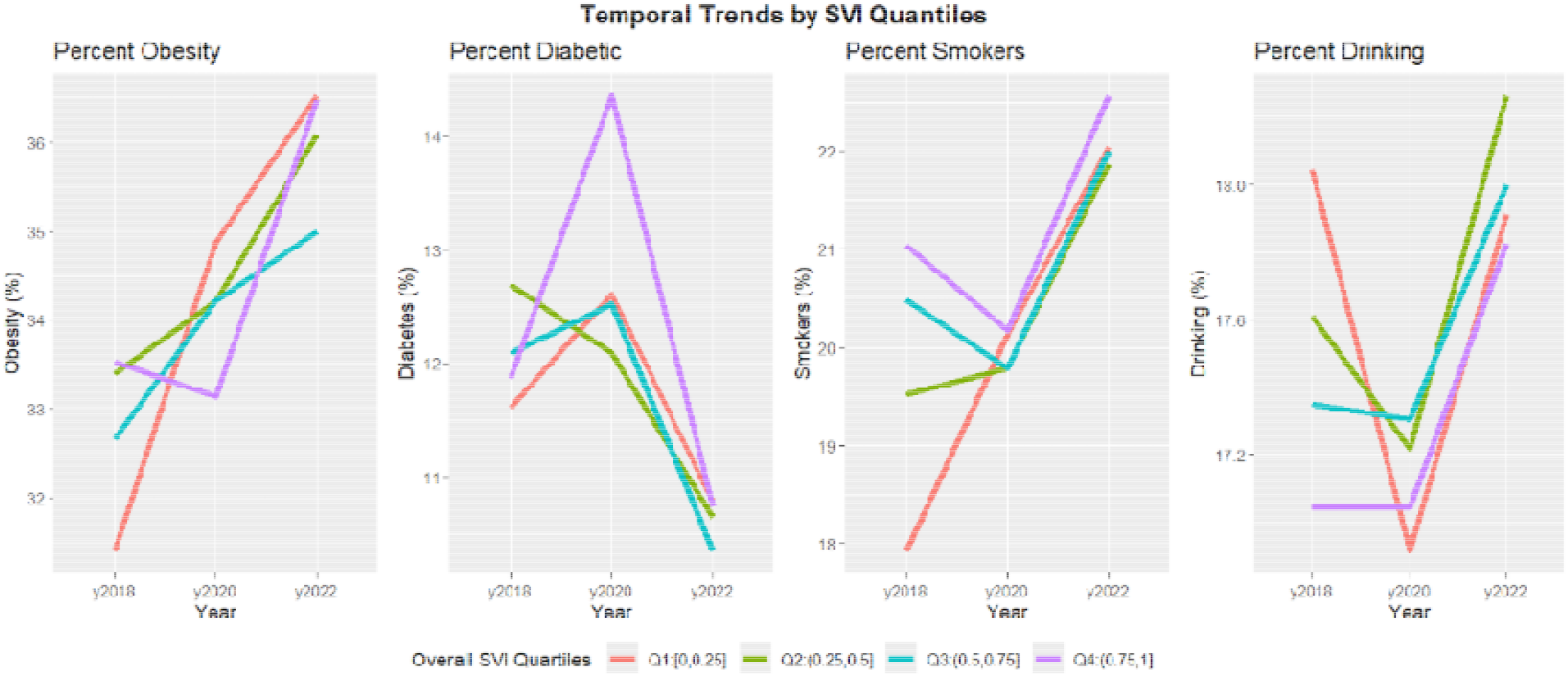
Temporal trends of comorbidities by SVI groups.

From **Figure 3**, we noticed that uninsured rates decreased from 2018 to 2020, saw an uptick from 2020 to 2022, but there was a net decrease in the percentage of uninsured individuals by 2022 for all quartiles except Q1, which saw a ~0.5% net increase. From 2018 to 2020, the number of mental health providers decreased in the SVI-Q4, but increased in the 1^st^, 2^nd^, and 3^rd^ SVI-Qs. Between 2020 and 2022, there was an increase in mental health providers across all SVI-Qs. Trends in primary care physicians were more variable. From 2018 to 2020, SVI-Q1 saw a dramatic decrease of a rate of ~10; SVI-Q2 experienced a dramatic increase of a rate of ~12.5; SVI-Q3 saw a moderate increase of a rate of ~1, and SVI-Q4 had a slight decrease of a rate of ~5. From 2020 to 2022, SVI-Q2 continued to increase, while the other SVI-Qs experienced declines in primary care physician numbers.

**Figure 3.**
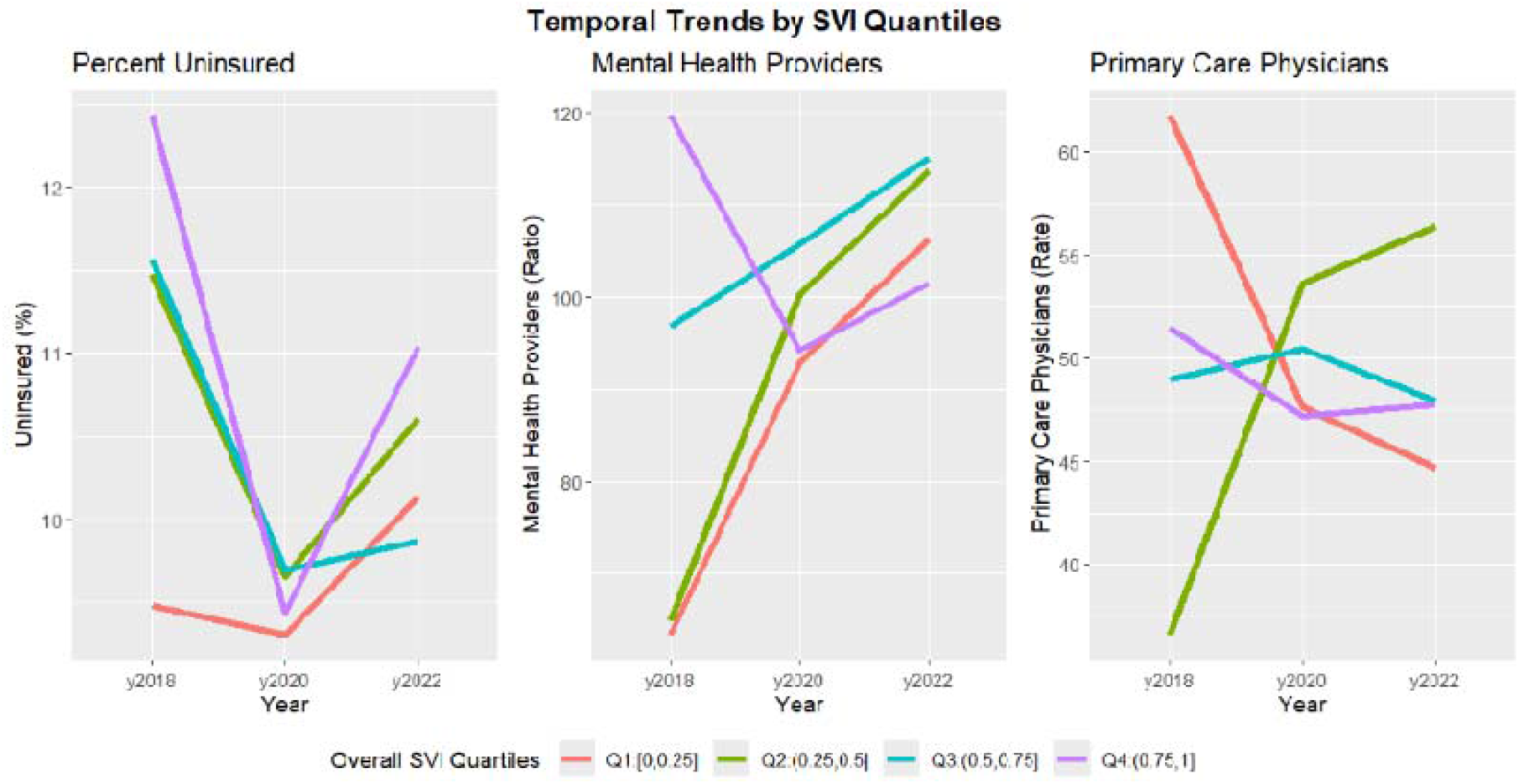
Temporal trends for variables representing access to healthcare and resources.

The correlation between obesity rate and access to exercise shows inconsistent and weak correlations across all SVI-Qs, with SVI-Q4 peaking positively at in 2020 before dropping sharply negatively in 2022 to about; SVI-Q3 remains consistently negative (**Figure 4**). In all SVI-Qs, correlation coefficients between obesity rate and physical inactivity increases steadily by 0.4, indicating a strengthening positive association; SVI-Q3 and SVI-Q4 show the strongest positive correlations by 2022. Correlation between obesity rate and food environmental index generally declines over time for all SVI-Qs. Here, SVI-Q3 started with the strongest positive correlation at in 2018 but decreased notably to by 2022; SVI-Q1 and SVI-Q2 showed increasingly negative correlations (from approximately to, for both quartiles); SVI-Q4 consistently showed little to no correlation.

**Figure 4.**
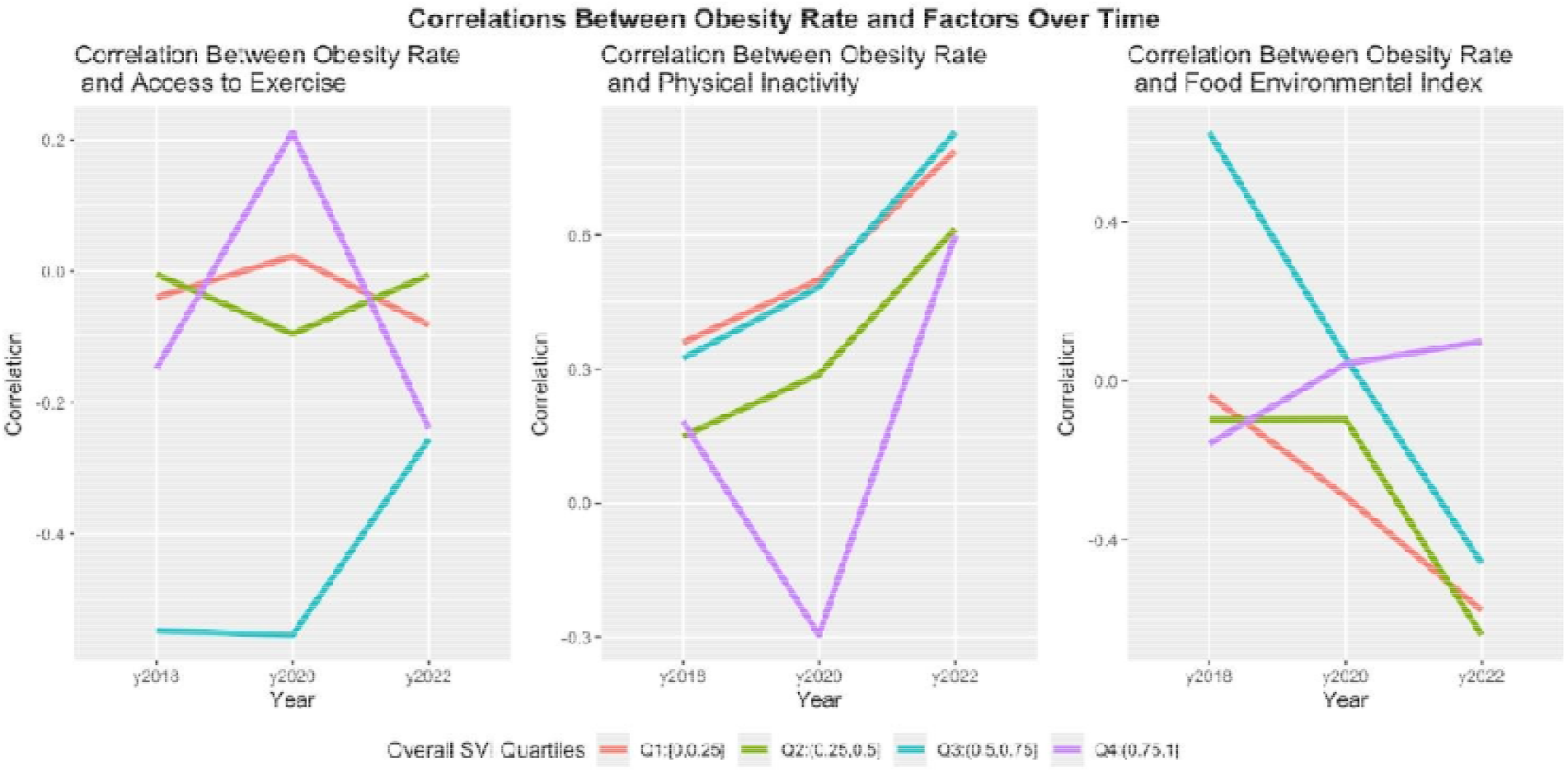
Correlations between obesity rate and factors over time.

From **Figures 5-7**, we noticed that across the three years, a persistent group of counties exceeded the 0.70 SVI threshold, such as counties Cass, Parke, Orange, and Saint Joseph. The number of counties with a higher-than-average percentage of premature death has slightly over time between 2018 and 2022, especially evident in the southwest region, but remains largely unchanged in central Indiana. Additionally, the number of counties classified as “Low SVI but High Premature Death” has grown steadily in the central region. Meanwhile, the number of “High SVI but Low Premature Death” counties remain small and largely unchanged.

**Figure 5.**
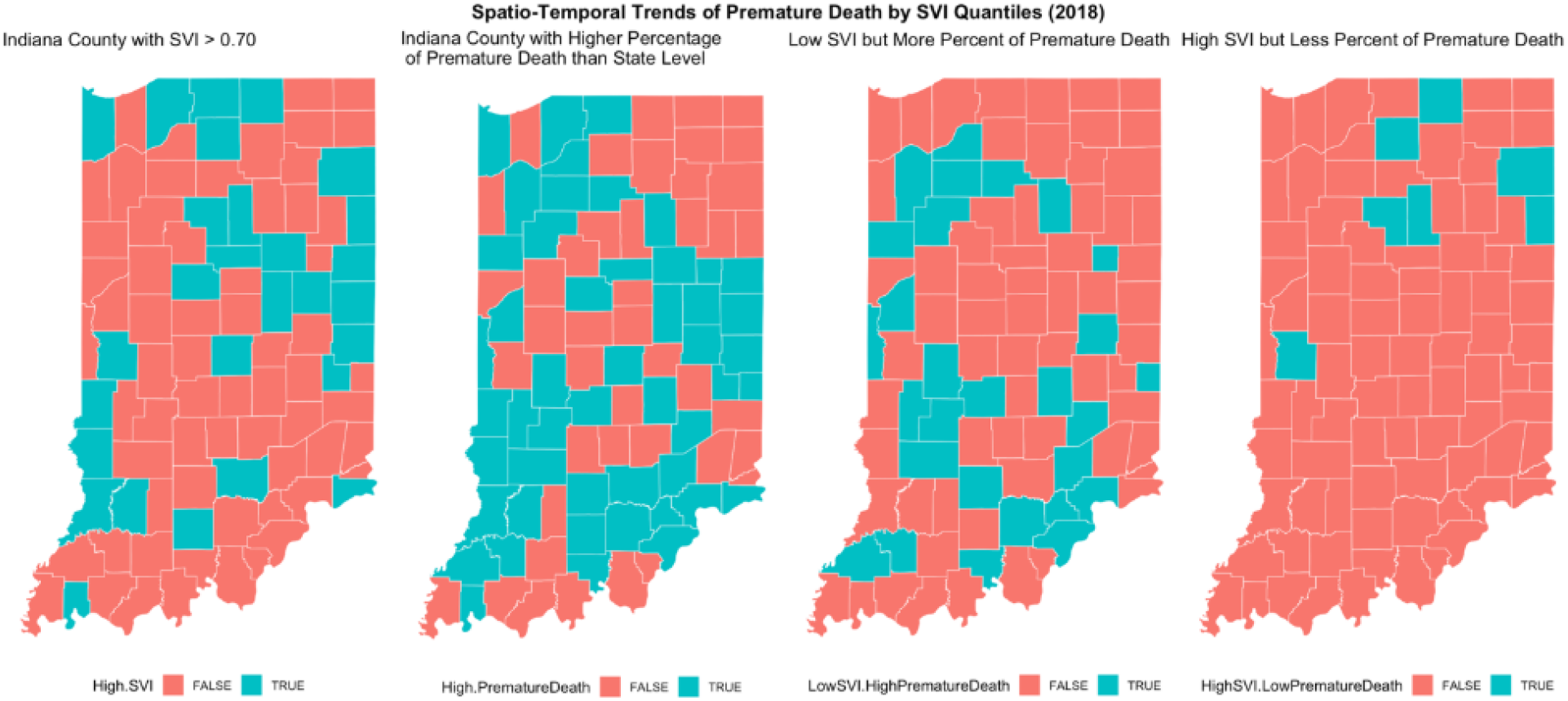
Spatiotemporal trends of premature death by SVI groups (2018).

**Figure 6.**
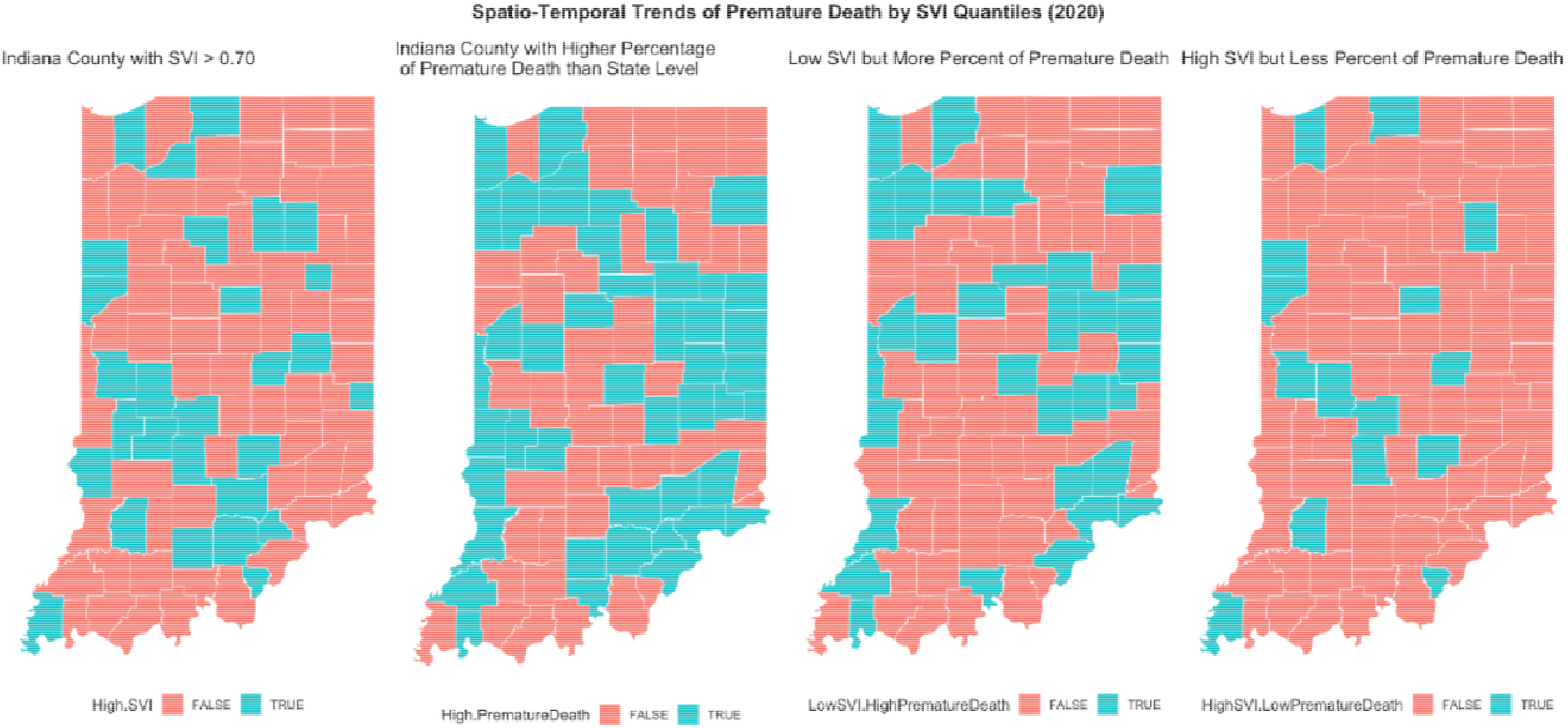
Spatiotemporal trends of premature death by SVI groups (2020).

**Figure 7.**
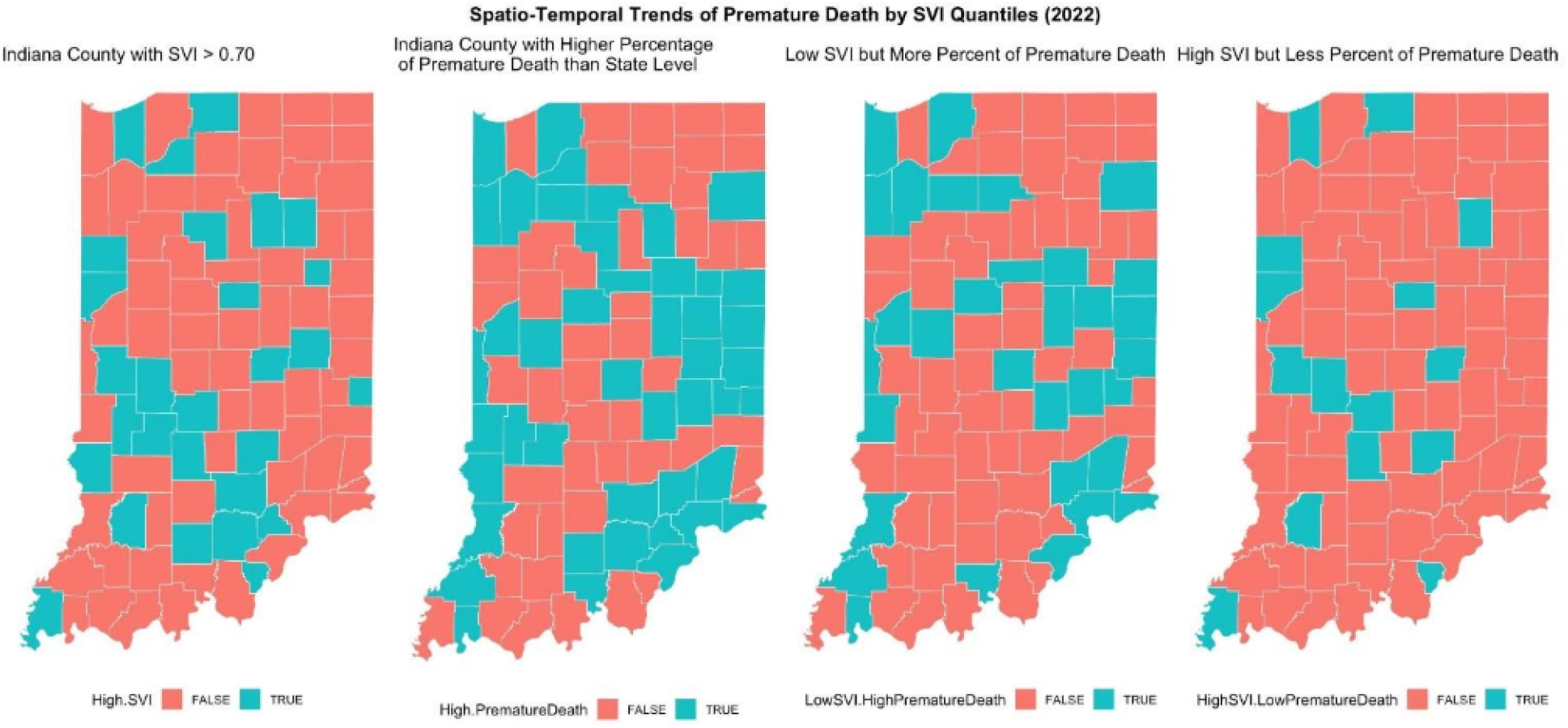
Spatiotemporal trends of premature death by SVI groups (2022).

Table 1. and **Figures 8-10** show the relationship between the different covariates with premature death after fitting year-specific NB regression models. From the NB regression model, an Incidence Rate Ratio (IRR) along with 95% confidence intervals and associated p-value are provided. IRR>1 indicate increased association while IRR<1 indicate decrease.

**Table 1.**
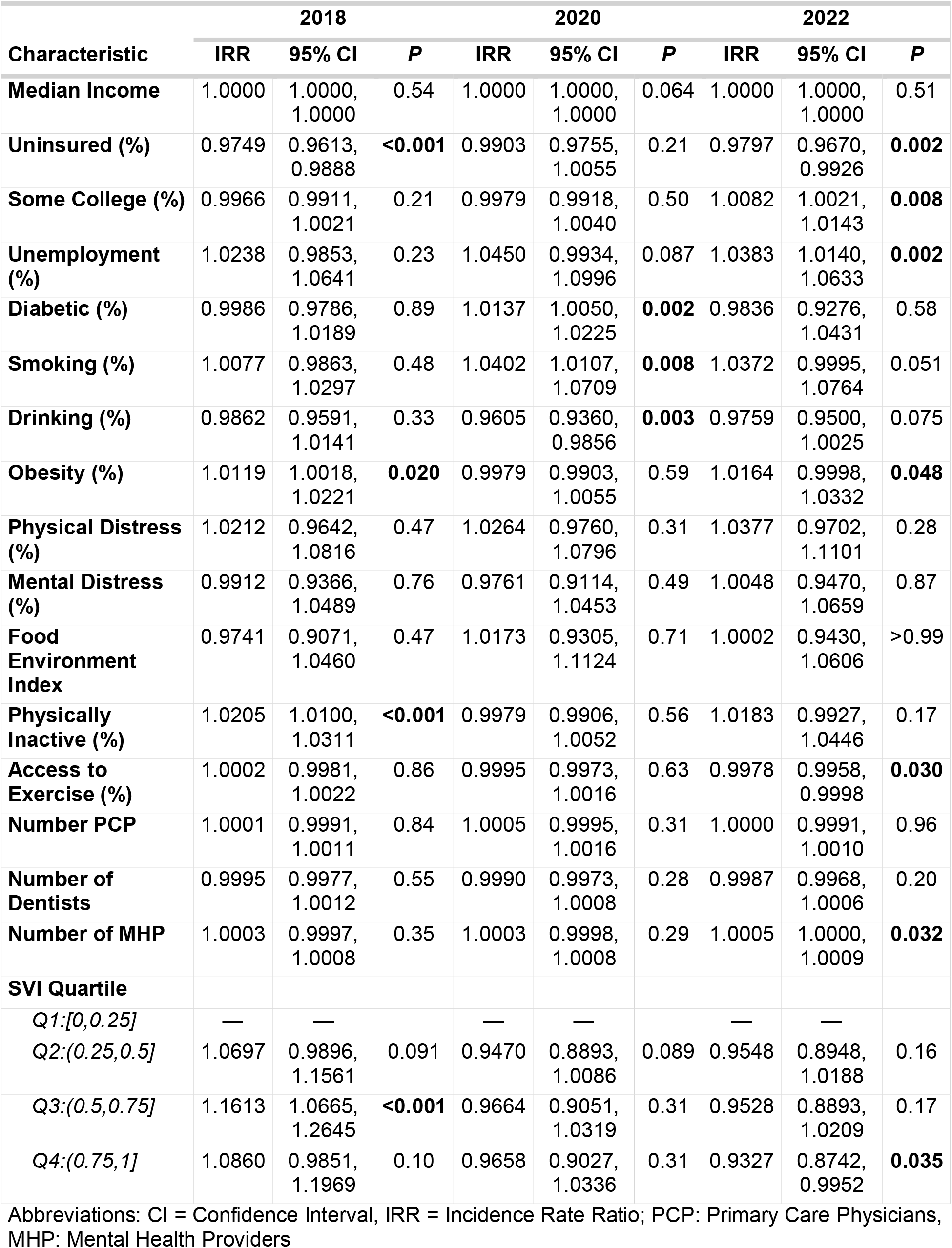
Results from Negative Binomial regression model with premature death as the outcome.

| Characteristic | 2018 |  |  | 2020 |  |  | 2022 |  |  |
| --- | --- | --- | --- | --- | --- | --- | --- | --- | --- |
|  | IRR | 95% CI | P | IRR | 95% CI | P | IRR | 95% CI | P |
| Median Income | 1.0000 | 1.0000, 1.0000 | 0.54 | 1.0000 | 1.0000, 1.0000 | 0.064 | 1.0000 | 1.0000, 1.0000 | 0.51 |
| Uninsured (%) | 0.9749 | 0.9613, 0.9888 | **<0.001** | 0.9903 | 0.9755, 1.0055 | 0.21 | 0.9797 | 0.9670, 0.9926 | **0.002** |
| Some College (%) | 0.9966 | 0.9911, 1.0021 | 0.21 | 0.9979 | 0.9918, 1.0040 | 0.50 | 1.0082 | 1.0021, 1.0143 | **0.008** |
| Unemployment (%) | 1.0238 | 0.9853, 1.0641 | 0.23 | 1.0450 | 0.9934, 1.0996 | 0.087 | 1.0383 | 1.0140, 1.0633 | **0.002** |
| Diabetic (%) | 0.9986 | 0.9786, 1.0189 | 0.89 | 1.0137 | 1.0050, 1.0225 | **0.002** | 0.9836 | 0.9276, 1.0431 | 0.58 |
| Smoking (%) | 1.0077 | 0.9863, 1.0297 | 0.48 | 1.0402 | 1.0107, 1.0709 | **0.008** | 1.0372 | 0.9995, 1.0764 | 0.051 |
| Drinking (%) | 0.9862 | 0.9591, 1.0141 | 0.33 | 0.9605 | 0.9360, 0.9856 | **0.003** | 0.9759 | 0.9500, 1.0025 | 0.075 |
| Obesity (%) | 1.0119 | 1.0018, 1.0221 | **0.020** | 0.9979 | 0.9903, 1.0055 | 0.59 | 1.0164 | 0.9998, 1.0332 | **0.048** |
| Physical Distress (%) | 1.0212 | 0.9642, 1.0816 | 0.47 | 1.0264 | 0.9760, 1.0796 | 0.31 | 1.0377 | 0.9702, 1.1101 | 0.28 |
| Mental Distress (%) | 0.9912 | 0.9366, 1.0489 | 0.76 | 0.9761 | 0.9114, 1.0453 | 0.49 | 1.0048 | 0.9470, 1.0659 | 0.87 |
| Food Environment Index | 0.9741 | 0.9071, 1.0460 | 0.47 | 1.0173 | 0.9305, 1.1124 | 0.71 | 1.0002 | 0.9430, 1.0606 | >0.99 |
| Physically Inactive (%) | 1.0205 | 1.0100, 1.0311 | **<0.001** | 0.9979 | 0.9906, 1.0052 | 0.56 | 1.0183 | 0.9927, 1.0446 | 0.17 |
| Access to Exercise (%) | 1.0002 | 0.9981, 1.0022 | 0.86 | 0.9995 | 0.9973, 1.0016 | 0.63 | 0.9978 | 0.9958, 0.9998 | **0.030** |
| Number PCP | 1.0001 | 0.9991, 1.0011 | 0.84 | 1.0005 | 0.9995, 1.0016 | 0.31 | 1.0000 | 0.9991, 1.0010 | 0.96 |
| Number of Dentists | 0.9995 | 0.9977, 1.0012 | 0.55 | 0.9990 | 0.9973, 1.0008 | 0.28 | 0.9987 | 0.9968, 1.0006 | 0.20 |
| Number of MHP | 1.0003 | 0.9997, 1.0008 | 0.35 | 1.0003 | 0.9998, 1.0008 | 0.29 | 1.0005 | 1.0000, 1.0009 | **0.032** |
| SVI Quartile |  |  |  |  |  |  |  |  |  |
| Q1:[0,0.25] | — | — |  | — | — |  | — | — |  |
| Q2:(0.25,0.5] | 1.0697 | 0.9896, 1.1561 | 0.091 | 0.9470 | 0.8893, 1.0086 | 0.089 | 0.9548 | 0.8948, 1.0188 | 0.16 |
| Q3:(0.5,0.75] | 1.1613 | 1.0665, 1.2645 | **<0.001** | 0.9664 | 0.9051, 1.0319 | 0.31 | 0.9528 | 0.8893, 1.0209 | 0.17 |
| Q4:(0.75,1] | 1.0860 | 0.9851, 1.1969 | 0.10 | 0.9658 | 0.9027, 1.0336 | 0.31 | 0.9327 | 0.8742, 0.9952 | **0.035** |

Abbreviations: CI = Confidence Interval, IRR = Incidence Rate Ratio; PCP: Primary Care Physicians, MHP: Mental Health Providers

**Figure 8.**
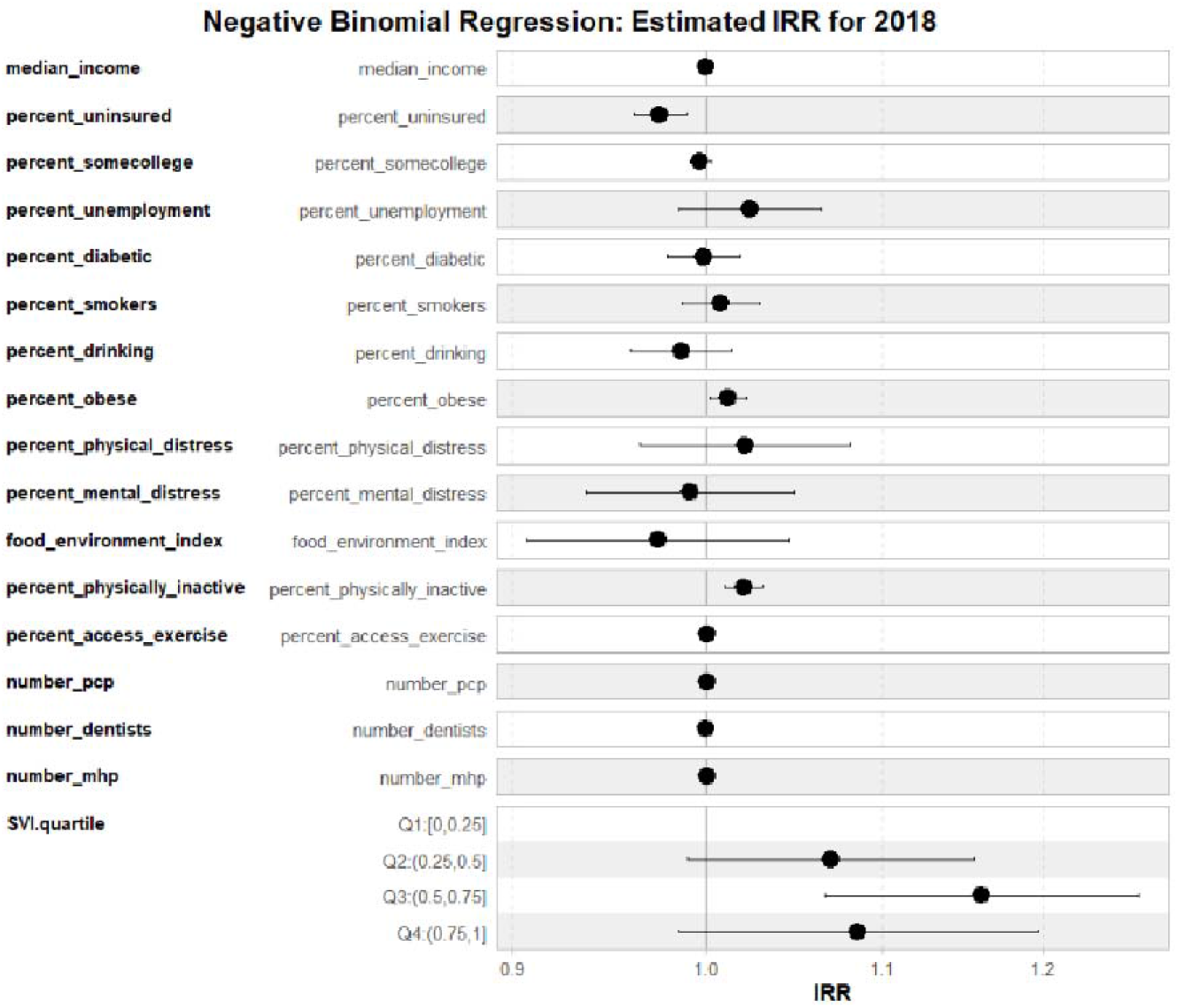
Results from Negative Binomial regression model. Estimated Incidence Rate Ratios (IRR) for 2018.

**Figure 9.**
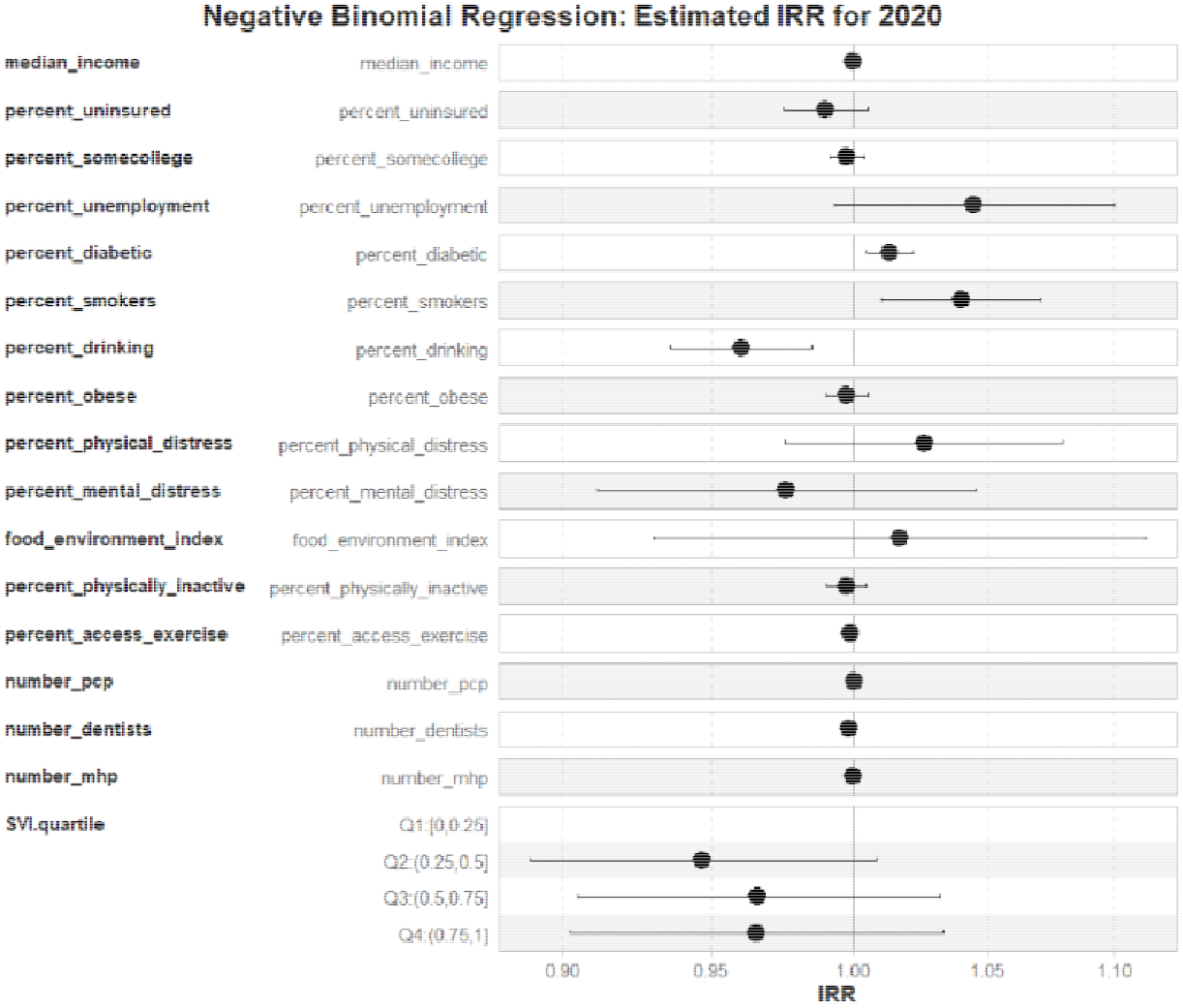
Results from Negative Binomial regression model. Estimated Incidence Rate Ratios (IRR) for 2020.

**Figure 10.**
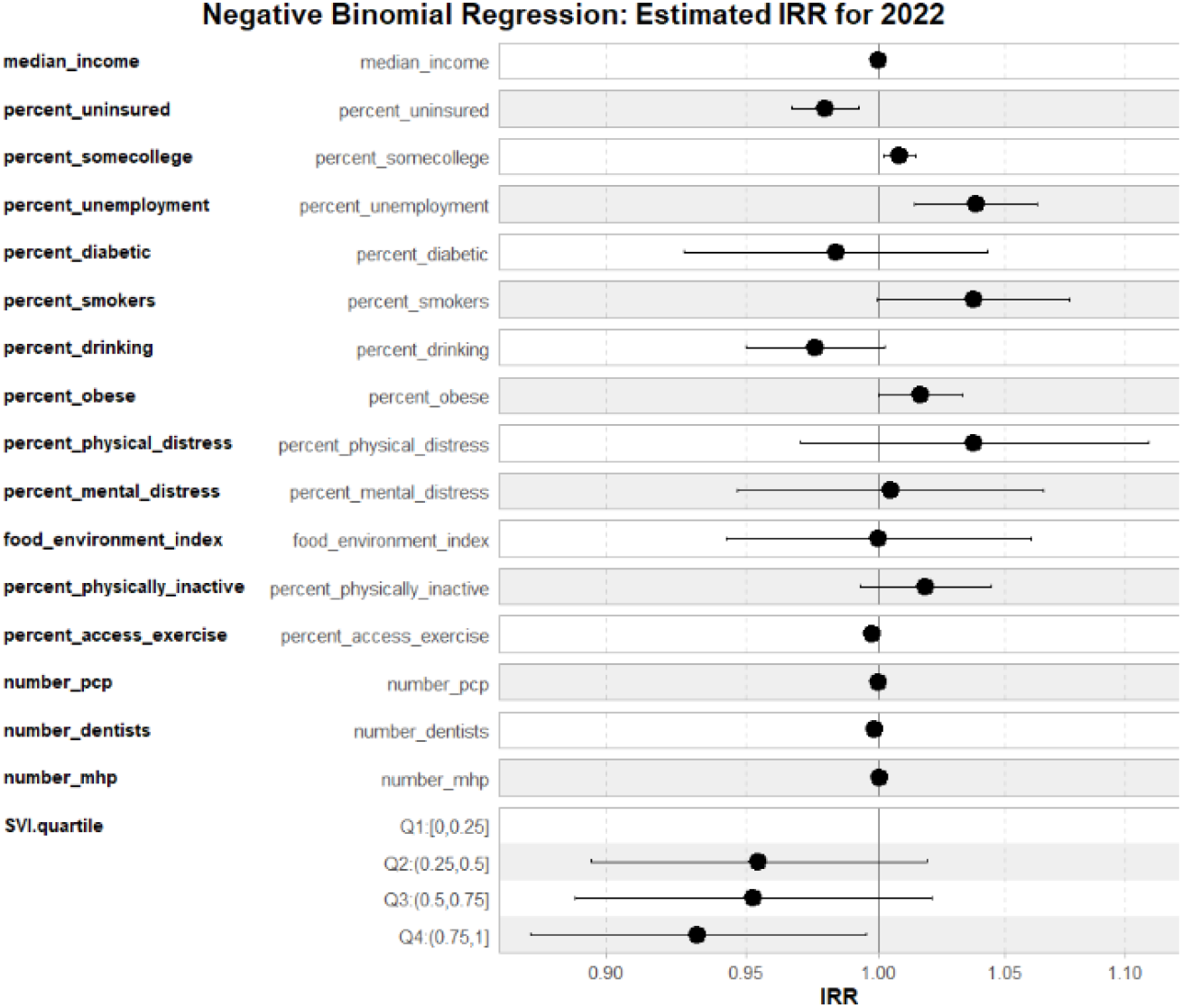
Results from Negative Binomial regression model. Estimated Incidence Rate Ratios (IRR) for 2022.

In 2018, we found that the percentage of uninsured (IRR: 0.9749; 95%CI: 0.9613 - 0.9888; p<0.001), percentage of obese (IRR: 1.0119; 95%CI: 1.0018 - 1.0221; p=0.02), and percentage of physically inactive individuals (IRR: 1.0205; 95%CI: 1.0100 - 1.0311; p<0.001) were statistically associated with premature death, with the last two associated with increased premature mortality.

In 2020, we found that the percentage of diabetics (IRR: 1.0137; 95%CI: 1.0050 - 1.0225; p=0.002), percentage of smoking (IRR: 1.0402; 95%CI: 1.0107 - 1.0709; p=0.008), and percentage of drinking (IRR: 0.9605; 95%CI: 0.9360 - 0.9856; p=0.003) were statistically associated with premature death, where drinking behavior was found to be associated with decreased outcome.

In 2022, we found that percentage of uninsured (IRR: 0.9797; 95%CI: .9670 - 0.9926; p=0.002), percentage of some college experience (IRR: 1.0082; 95%CI: 1.0021 - 1.0143; p=0.008), percentage of unemployment (IRR: 1.0383; 95%CI: 1.0140 - 1.0633; p=0.002), percentage of obese (IRR: 1.0164; 95%CI: 0.9998 - 1.0332; p=0.048), percentage of access to exercise (IRR: 0.9978; 95%CI: 0.9958 - 0.9998; p=0.030), and number of mental health providers (IRR: 1.005; 95%CI: 1.0000 - 1.0009; p=0.032) were statistically associated with premature death.

## Discussion

This study explored trends in health outcomes across Indiana counties with varying levels of social vulnerability before, during, and after the COVID-19 pandemic. Here we presented some insights into how the COVID-19 pandemic may have amplified existing health disparities in socially vulnerable counties in the state of Indiana. Our findings align with previous work showing that structural inequities, including occupational risk, income, access to care, and neighborhood environments, exacerbate the health consequences of public health crises.^5^ The trends observed across physical health, mental health, obesity, insurance coverage, and premature death suggest that the pandemic intensified existing vulnerabilities.

National data from the Behavioral Risk Factor Surveillance System revealed that average BMI among US adults increased and obesity prevalence rose during the pandemic.^15^ Similarly, the data used in this study demonstrated continued increases in adult obesity rates even after 2020, suggesting that the pandemic’s impact on weight gain was both widespread and persistent. The rise in alcohol consumption in Indiana after 2020 was also an observed national trend.^16^ Since alcohol is calorie-dense and can interfere with fat metabolism,^17^ both the state and national trends show that increased alcohol consumption likely contributed to higher obesity rates.^16^ However, while national data saw a decline in smoking rates,^16^ our Figure 2 suggests a state-wide increase in smoking among all SVI groups. The observed difference in smoking trends may be partially attributable to the timing and implementation of Indiana’s Tobacco 21 law,^18^ which took effect in July 2020 and raised the legal age for purchasing tobacco and vapor products to 21 years old. National declines in smoking were likely influenced by stronger enforcement and broader awareness, whereas Indiana’s rural populations may have been slower to adapt to the law or maintained higher tobacco use due to economic stressors and cultural norms.^19^ Additionally, while national declines in smoking are sometimes linked to modest weight gain due to nicotine withdrawal, Indiana’s slower decrease or temporary increase in smoking may have mitigated that indirect contribution to BMI growth.

A clear increase in poor physical health days was observed across the counties in Indiana, particularly in those with higher SVI scores. One plausible explanation is the reduced opportunities for physical activity during the pandemic. Socially vulnerable populations often face structural barriers to exercise, including limited access to safe outdoor spaces and recreational facilities.^20^ Pandemic-related closures further compounded this, resulting in prolonged physical inactivity.^21^ Physical inactivity has been recorded as strongly associated with worse self-reported health, obesity, and chronic disease,^22^ which helps explain the simultaneous rise in poor physical health days and obesity documented in this study. In line with this, prior studies have reported that low-income and minority populations were disproportionately impacted by restricted exercise opportunities during the pandemic.^4^

The rise in poor mental health days mirrors national and international findings that the pandemic had significant psychological consequences.^20^ Our results may reflect the combined stressors of social isolation, economic insecurity, and family disruption. For children and adolescents, school closures and increased screen time have been linked to higher rates of anxiety, depression, and behavioral problems. These factors help to possibly explain why poor mental health days increased even though mental health services expanded post-2020. The persistence of this trend underscores the mismatch between service availability and accessibility, particularly in socially vulnerable counties, where stigma, financial barriers, and shortages of providers may limit uptake of care.

Obesity trends observed in Figure 2 provide further evidence of the pandemic’s indirect health effects. Rates of obesity rose sharply across all SVI quartiles, consistent with reports from other states.^4^ Several factors likely contributed to this increase of obesity rates, including reduced physical activity due to stay-at-home orders, increased consumption of calorie-dense foods during lockdowns, and elevated stress levels driving unhealthy eating behaviors.^4^ Interestingly, diabetes prevalence declined slightly in the same period. This apparent contradiction could be due to under-diagnosis during the pandemic, as fewer individuals sought preventive care and routine screenings, rather than a true reduction in disease burden.^20^ Thus, the sharp rise in obesity paired with a decrease in diagnosed diabetes may reflect healthcare disruptions rather than genuine improvements in metabolic health.

The negative correlation between obesity and the food environment index in Figure 4 highlights the role of environmental determinants. Food deserts, areas with poor access to affordable, nutritious food, are more common in socially vulnerable communities and contribute to unhealthy dietary patterns.^4^ Pandemic-related disruptions in food supply chains and income loss may have exacerbated reliance on cheaper, calorie-dense foods, further fueling obesity trends. Similarly, the positive correlation between obesity and poor physical health underscores the reinforcing cycle between inactivity, poor health, and weight gain.

Interestingly, while a national study documented a rise in exercise participation and increase in sleep duration,^23^ obesity still rose. This mirrors the paradox seen in Figure 4, where increased access to exercise opportunities after 2020 did not correspond to reduced obesity rates. Both findings may imply that moderate improvements in exercise and rest were insufficient to counterbalance other behavioral and metabolic risk factors, such as the increased caloric intake and stress-related eating.

Insurance coverage trends also shed light on amplified health inequities. Figure 3 shows that uninsurance rates rose between 2020 and 2022 in counties within the highest SVI quartile. Job losses during the pandemic disproportionately affected low-income and minority workers, many of whom rely on employer-sponsored insurance.^20^ Losing coverage likely delays care-seeking behaviors, contributing to more severe illness and potentially higher mortality rates.^24^ These findings align with national evidence that uninsured individuals faced worse outcomes during the pandemic, both for COVID-19 and other chronic conditions.^25^

Finally, premature death patterns from 2018 to 2020 showed consistently higher rates in counties with low SVI, while some high-SVI counties experienced lower rates. Although unexpected, this may reflect urban–rural dynamics. High-SVI counties in Indiana are more likely to be urban, and urban populations often skew younger, which could reduce premature mortality despite greater social vulnerability. Conversely, rural low-SVI counties may have older populations and less access to healthcare infrastructure, driving higher premature death rates.^26^ Similar studies have highlighted the paradox of rural disadvantage, where low social vulnerability scores may not capture healthcare scarcity or aging demographics that elevate mortality risk.^27^

Taken together, these results suggest that the health disparities observed during the pandemic were not isolated outcomes, but the product of multiple potential and reinforcing causal pathways. Occupational exposure reduced exercise opportunities, disruptions in healthcare, food environment limitations, and structural barriers to care all contributed to the amplified burden in socially vulnerable counties. Our findings are consistent with broader research showing that socially vulnerable populations are disproportionately impacted during public health crises, but they also highlight the importance of contextual factors, such as urbanicity and healthcare infrastructure, that may mediate these outcomes.^20^ Targeted interventions addressing these underlying determinants, including improved access to exercise opportunities, food support, mental health services, and health insurance coverage, will be critical for mitigating long-term impacts in vulnerable communities.

The study had some limitations. Since data on state laws and policies were not available in the CHR&R datasets used for this study, particularly for the selected years, we were unable to incorporate these important factors into the models as covariate. We plan to consider these factors in future analyses as data of federal and state laws and policies become available within county-level. Future works will consider the exploration of the interactions between a subset of the candidate covariates, and a stratified analysis. We also plan to consider a formal, causal inference framework into our future work to capture potential cause-and-effect results rather than just associations. Future analyses should also investigate how pandemic-era behavioral changes interacted with local policy measures and food environment quality. Incorporating individual-level data could clarify causal links between these behaviors and long-term obesity and mental health outcomes. Further, including post-2022 data would help determine whether health outcomes trends stabilize, worsen, or improve years after the pandemic. Understanding how policy enforcement, food access, and economic recovery influence these patterns will be critical for developing targeted interventions that reduce obesity, mental health issues, and related chronic disease risks in Indiana’s most vulnerable communities and beyond.

## Conclusion

This study highlights how social and economic vulnerability continue to shape health outcomes across Indiana counties. The analysis revealed that counties with higher social vulnerability experienced greater increases in obesity, poorer mental health, and higher premature death rates. Despite modest improvements in healthcare access and the growth of mental health services after 2020, disparities in overall health outcomes persisted and, in many cases, widened. Findings from this work suggest that structural barriers, limited access to exercise opportunities, inadequate food environments, and uneven healthcare coverage, may have intensified the pandemic’s effects, particularly in rural and low-income communities. While mental health providers increased post-pandemic, the rise in reported poor mental health days indicates that availability alone is insufficient without equitable distribution and accessibility of services. The results underscore the urgent need for targeted, data-driven interventions that strengthen public health infrastructure and address upstream social determinants of health. Expanding access to recreational facilities, improving food security, and supporting sustainable mental health care delivery in high-vulnerability areas will be critical to narrowing these health gaps. By integrating social determinants into statewide public health planning, Indiana can move toward more resilient, equitable communities better prepared for future public health crises.

## Data Availability Statement

All data produced are available online at the County Health Rankings and Roadmaps database, obtained from the University of Wisconsin Population Health Institute’s public website, and the Social Vulnerability Index (SVI) data files from the Centers for Disease Control and Prevention website.

## Acknowledgements

We would like to thank our mentor, Dr. Pabon-Rodriguez, and his PhD student, Plamena Powla, for graciously providing guidance throughout the research process and supporting the development of this study.

## Authors

**Kelli Wang** is a high school senior, who will be attending Northwestern University in the Fall 2026. She plans to study neuroscience with a minor in music cognition on the pre-medical track. Her interests are in public health, cancer research, and interdisciplinary applications of neuroscience.

**Precious Olaniyan** was a high school senior while completing this research project in Summer 2025. She joined Northwestern University in Fall 2025 majoring in Biology and Global Health. She is interested in Pediatrics and Internal Medicine and is deeply committed to addressing health disparities through equitable, patient-centered care.

